# Genomic relationships across psychiatric disorders including substance use disorders

**DOI:** 10.1101/2020.06.08.20125732

**Authors:** Abdel Abdellaoui, Dirk J.A. Smit, Wim van den Brink, Damiaan Denys, Karin J.H. Verweij

## Abstract

**Background and aims:** A recent study investigated the genetic associations and latent genetic structure among eight psychiatric disorders using findings from genome-wide association studies (GWASs). No data from substance use disorders were included, while these represent an important category of mental disorders and could influence the latent genetic structure. We extended the original paper by recreating the genetic relationship matrix, graph, and latent genetic factor structure, including additional data from substance use disorders.

**Methods:** We used GWAS summary statistics of 11 psychiatric disorders, including alcohol dependence, nicotine dependence, and cannabis use disorder. We estimated genetic correlations between all traits in Linkage Disequilibrium-score regression. A graph was created to illustrate the network of genetic correlations. We then used the genetic relationships to model a latent genetic factor structure.

**Results:** Alcohol and nicotine dependence showed significant genetic correlations with several other psychiatric disorders, including ADHD, schizophrenia, and major depression. Cannabis use disorder was only significantly associated with ADHD. The addition of substance use disorders resulted in some changes in the latent structure of the factor model when compared to the original model including eight disorders. All substance use disorders contributed mostly to Factor 3, a heterogeneous factor with also loadings from ADHD, major depression, Autism Spectrum disorders, and Tourette Syndrome.

**Conclusions:** Alcohol and nicotine dependence show widespread genetic correlations with other psychiatric disorders. Including substance use disorders in the factor analysis results in some changes in the underlying genetic factor structure. Given the instability of such models, identified structures should be interpreted with caution.

## Introduction

Last year, Lee et al. (1) published a paper in Cell in which they combined genome-wide association study (GWAS) data from eight psychiatric disorders - anorexia nervosa (AN), attention-deficit/hyperactivity disorder (ADHD), autism spectrum disorder (ASD), bipolar disorder (BIP), major depression (MD), obsessive-compulsive disorder (OCD), schizophrenia (SCZ), and Tourette syndrome (TS) – to identify novel pleiotropic genes and a latent genetic structure underlying these eight disorders. Using exploratory factor-analyses, and confirmed by hierarchical clustering analyses, they identified three (correlated) factors explaining approximately half of the genetic variation in these eight disorders. The first factor consisted primarily of disorders characterized by compulsive behaviours (AN, OCD, and TS), the second by mood and psychotic disorders (MD, BIP, and SCZ), and the third by early-onset neurodevelopmental disorders (ASD, ADHD, and TS) as well as MD. The authors conclude that their findings have important implications for psychiatric nosology, i.e. the classification of psychiatric disorders.

Unfortunately, the authors did not include any data from substance abuse or dependence GWASs in their analyses, while substance use disorders form an important category of mental disorders. Substance use disorders are highly impairing, causing great harm to the individual, their family and friends, and to the society as a whole (2). There is evidence for substantial phenotypic and genetic correlations between substance use disorders and other psychiatric disorders.(3) The decision to not include substance use disorders in the analyses is a potential caveat, in particular because the investigated traits now most of the lack externalising disorders. The limited inclusion of traits may, therefore, have biased the identified latent structure of disorders.

We extended the analyses by Lee et al. (1) by including additional data from three GWASs on substance use disorders: alcohol, nicotine, and cannabis dependence. We investigated the genetic relationships between these 11 traits by recreating the genetic relationship matrix and graph and by producing a new latent genetic factor structure.

## Methods

We used data from the same eight GWAS studies of psychiatric disorders conducted by the Psychiatric Genomics Consortium: attention-deficit/hyperactivity disorder (ADHD, 4), anorexia nervosa (AN, 5), autism spectrum disorder (ASD, 6), bipolar disorder (BIP, 7), major depression (MD, 8), obsessive-compulsive disorder (OCD, 9), schizophrenia (SCZ, 10), and Tourette syndrome (TS, 11). For MD we only had access to data excluding 23andMe, resulting in smaller sample than Lee et al. (1) used. In addition, we included GWAS data from three GWASs for substance use disorders: alcohol dependence (AD; DSM-IV-diagnosis, 12), cannabis use disorder (CUD, ICD diagnosis, 13), and nicotine dependence (ND, Fagerström Test for Nicotine Dependence; dividing the sample in: mild (scores 0–3), moderate (scores 4–6), and severe (scores 7–10), 14). See Table 1 for sample sizes and SNP-based heritabilities for each disorder, and the original publications for detailed sample descriptions.

**Table 1:** Sample sizes and SNP-based heritabilities as computed using LDSC regression.

| **Psychiatric Disorder** | **Cases** | **Controls** | **Total** | **Effective Sample Size** | **SNP-based $h^2$ (SE)** |
| --- | --- | --- | --- | --- | --- |
| ADHD (4) | 19,099 | 34,194 | 53,293 | 49,017.41 | .26 (.01) |
| AN (5) | 3,495 | 10,982 | 14,477 | 10,604.98 | .33 (.05) |
| ASD (6) | 18,382 | 27,969 | 46,351 | 44,368.07 | .20 (.02) |
| BIP (7) | 20,353 | 31,358 | 51,710 | 49,368.94 | .36 (.02) |
| MD (8) | 59,851 | 113,154 | 173,005 | 156,582.3 | .08 (.01) |
| OCD (9) | 2,688 | 7,037 | 9,725 | 7,780.136 | .40 (.06) |
| SCZ (10) | 36,989 | 11,3075 | 150,064 | 111,486.6 | .32 (.01) |
| TS (11) | 4,819 | 9,488 | 14,307 | 12,783.3 | .41 (.05) |
| AD (12) | 11,569 | 34,999 | 46,568 | 34,779.54 | .10 (.02) |
| CUD (13) | 2,387 | 48,985 | 51,372 | 9,104.352 | .16 (.06) |
| ND (14) | NA | NA | 28,677 | 28,677 | .12 (.02) |

*ADHD = attention-deficit/hyperactivity disorder; AN = anorexia nervosa; ASD = autism spectrum disorder; BIP = bipolar disorder; MD = major depression; OCD = obsessive-compulsive disorder; SCZ = schizophrenia; TS = Tourette syndrome; AD = alcohol dependence; CUD = cannabis use disorder; ND = nicotine dependence.*

We estimated bivariate genetic correlations between all pairs of traits using LD-score (LDSC) regression (15). The genetic correlations are based on the estimated slopes from the regressions of the product of z-scores from two sets of GWAS summary statistics on the linkage disequilibrium (LD) score and represent the genetic covariation between two traits based on all polygenic effects captured by the genome-wide SNPs. The genome-wide LD patterns used by these methods were estimated in the European populations included in the HapMap 3 reference panel (16, 17). The LDSC regression analyses were performed using the 1,290,028 genome-wide HapMap SNPs used in the original LD score regression studies (16, 17).

A graph was plotted with the absolute genetic correlations as strength parameter for the edges (connections) using R version 3.5.1 (18) and R package igraph (19). Layout followed the Fruchterman-Reingold algorithm. Non-significant weights (*p*>0.05 after Bonferroni correction) were removed. Centrality of each trait was assessed with eigenvector centrality (20), and reflects whether the trait has strong genetic overlap with traits that themselves have strong overlap with other traits.

We used the genetic correlations derived from the genome-wide genetic data to model the joint genetic architecture of the 11 psychiatric disorders, closely following the methods reported by Lee et al. (1). First, an exploratory factor analysis (EFA) was performed with a predefined three-factor extraction and promax rotation. This determined the factor structure for the subsequent confirmatory factor analysis (CFA), which we performed in Genomic Structural Equation Modelling (Genomic SEM, 21). Three correlated factors were created, including only paths to traits with path loadings ≥.20 in the EFA. The diagonally weighted least squares (DWLS) estimator was used. The full genetic relationship matrix was used as input. Comparative fit index (CFI) and standardised root-mean-square residual (SRMR) were used as model fit parameters. Secondly, we employed the same procedure adding a fourth factor, to establish whether substance use disorders would form a separate factor from the other psychiatric disorders.

## Results

The bivariate genetic correlations between all pairs of traits are presented in Figure 1A and the exact estimates and standard errors are presented in Supplementary Table 1. Of the initial 8 psychiatric disorders, ADHD and MD showed the highest genetic correlations with the substance use disorders, with ADHD showing significant genetic correlations with all three substance use disorders (ADHD-AD: *r*_g_=.46, *p*=3.8×10^−7^, ADHD-ND: *r*_g_=.54, *p*=3.3×10^−13^, ADHD-CUD: *r*_g_=.35, *p*=7.1×10^−4^) and major depression only with alcohol and nicotine dependence (MD-AD: *r*_g_=.52, *p*=1.3×10^−10^, MD-ND: *r*_g_=.45, *p*=9.4×10^−12^). Both alcohol and nicotine dependence showed significant genetic correlations with four psychiatric disorders, whereas cannabis use disorder showed significant genetic correlations only with ADHD.

**Figure 1.**
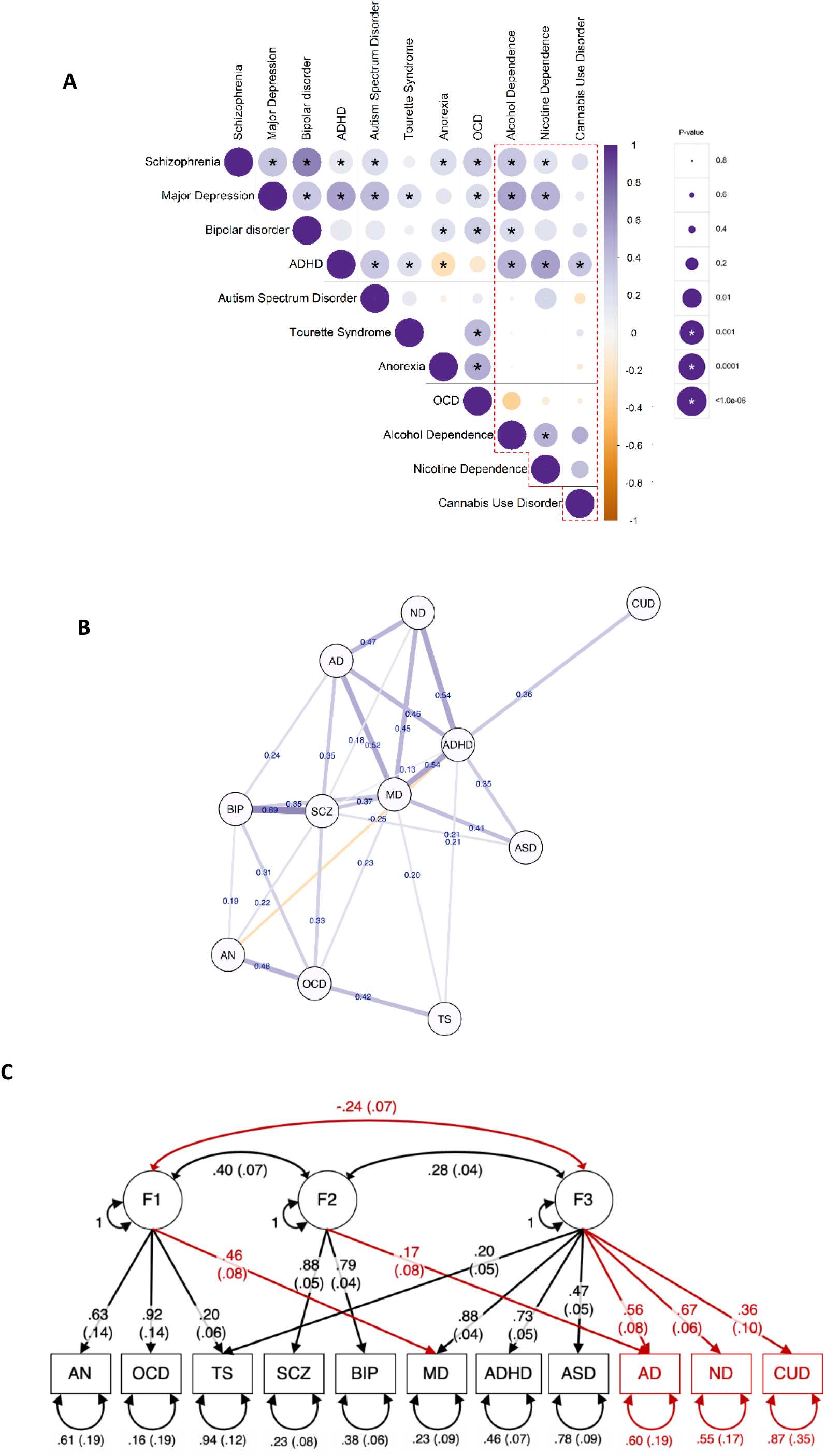
Genetic relationships between eleven psychiatric disorders. **A**. SNP-based genetic correlations (r_g_) between 11 psychiatric disorders (including 3 substance use disorders) using LD-Score regression. The size of the circles represents the significance of the *p*-values, whereas the colour represents the magnitude of the genetic correlation (darker = stronger). Asterisks indicate statistical significance after Bonferroni correction for multiple testing (.05/55 = .0009). **B**. Graphical representation of the SNP-based genetic correlations between 11 psychiatric disorders. Each vertex (node) represents a disorder. Edge strength was set to the value of the genetic correlation between the traits. Only edges representing significant genetic correlations after Bonferroni correction are shown. The width and colouring of the edges represent the strength of the genetic correlation (absolute values of r_g_). Positive genetic correlations are presented in blue, negative correlations in orange. **C**. Results of the genetic factor analysis. Each latent genetic factor represents shared genetic variance across the psychiatric disorders that load on it. One-headed arrows connecting the latent genetic factors to the individual disorders represent standardized loadings, which can be interpreted as coefficients from a regression of the true genetic liability for the disorder on the common factor. Two-headed arrows between the three latent factors represent their correlations. Two-headed arrows below the individual psychiatric disorders represent residual genetic variance not explained by the latent factor(s). Standardized parameter estimates are shown with their standard errors in parentheses. In red we show qualitative changes between the results of our genetic factor analyses with 11 psychiatric disorders and those of the original analysis by Lee et al.(1) including eight disorders.

Figure 1B shows the genetic relationship graph, displaying the significant SNP-based genetic correlations as determined in LD-score regression. Similar to the plot by Lee et al. (1), major depression takes a central place in the graph with many strong genetic correlations with other traits, now also including alcohol and nicotine dependence. The graph illustrates that ADHD also takes in a central place, with additional significant and strong genetic correlations with all three substance use disorders. The central positions of major depression and ADHD were reflected in the fact that they showed the highest eigenvector centrality scores (0.47 and 0.40, respectively).

To verify whether our approach matched those reported before, we replicated the EFA for eight disorders from Lee et al.(1) without substantial differences, with the same good model fit (CFI= 0.97, SRMR= 0.08) and roughly identical path loadings. Adding the three substance use disorders also yielded a good CFI fit index (0.95), and reasonable SRMR (0.10). The three correlated latent factors together explained 51% of the genetic variation in the 11 psychiatric disorders. Figure 1C shows the results of the CFA as obtained from Genomic SEM, including paths with path loadings above a threshold of ≥0.20 in the EFA. Qualitative changes in the model when compared to the original model by Lee et al. are shown in red. The factor loadings largely remained the same, however, some notable changes appeared compared to the previous analysis. Similar to the results by Lee et al. (1), Factor 1 consists primarily of disorders characterized by compulsive/perfectionistic behaviours (AN, OCD, and TS), but - in contrast to their results - we now also find a significant and substantial loading of MD. Our second factor is characterized by psychotic disorders (BIP and SCZ) and alcohol dependence, whereas the loading of major depression is no longer significant. Our third factor is now characterised by a large number of disorders, including MD, ADHD, ASD, and TS, and all three substance use disorders. We now also found a significant negative correlation between Factors 1 and 3.

Adding a fourth factor did not converge. To aid convergence, we forced the removal of additional path loadings by increasing the threshold from 0.20 to 0.35, which forced a single loading for each disorder. This model converged but yielded a poor fit (CFI<0; RMSR>1), strongly suggesting that a four-factor model is not adequate.

## Discussion

We extended analyses of a recent study investigating the genetic associations and underlying genetic factor structure among eight psychiatric disorders, by including three substance use disorders. We show that alcohol and nicotine dependence are significantly genetically correlated with several of the other mental health disorders, including ADHD, schizophrenia, and major depression. Cannabis use disorder is only significantly associated with ADHD, but this may be due in part to the limited power of the cannabis use disorder GWAS.

The addition of the substance use disorders to the genetic factor analysis provides additional information on the psychiatric three-factor genetic structure identified by Lee et al. (1). Factor 1 (AN, OCD, TS, and MD) and 2 (BIP, SCZ, AD) remained quite similar, but MD now loads on Factor 1, whereas in the original model MD loaded on Factor 2 (although MD still contributes to Factor 2 through the inter-factor correlations). In addition, alcohol dependence now also significantly contributes to Factor 2. Factor 3 is characterised by its large number of disorders, including all the disorders identified in the original structure (TS, MD, ADHD, ASD) plus all three substance use disorders. A four factor model did not fit the data well, suggesting that substance use disorders do not form a latent factor separate from the previously identified factors or that substance use disorders together with ADHD form a simple externalizing genetic trait. Lastly, we now observe a negative correlation between Factor 1 and 3, which was not the case in the original model by Lee et al.

While Lee et al. referred to their third factor as consisting mainly of early-onset neurodevelopmental disorders (ASD, ADHD, and TS), the new factor 3 is more heterogeneous. It includes four externalising disorders (ADHD, AD, ND, CUD) as well as one internalizing disorder (MD) and other less clearly positioned disorders (ASD and TS). Major depression, is a very common mental health disorder and associated with the broad range of psychiatric problems, so its contribution to multiple factors –including this factor-is not particularly surprising.

The switch of major depression contributing significantly to Factor 2 in Lee et al.’s factor model and to Factor 1 in our factor model highlights the sensitivity of such models to the input data. Aside from the additional input data for substance use traits, the input data for major depression in our model was a subset of the data used by Lee et al (i.e. their meta-analytic sample without 23andMe data), which could also explain some of the differences in the model. Updating source data (when larger GWASs come out) and adding additional disorders will likely result in additional changes or refinements in the underlying genetic factor structure. Larger GWASs will result in more power to detect significant genetic associations, changing the genetic correlation matrix and the genetic relationship graph. For example, the cannabis use disorder GWAS has substantial genetic correlations with some other traits (see Supplementary Table 1), but these were not significant likely due to the limited sample size of the GWAS. Furthermore, it is likely that including GWAS data on other diagnostic data, such as personality disorders, sleep-wake disorders, panic disorders, sexual dysfunction, neurological disorders, or other disorders not included in the current analyses, would lead to further changes in the underlying structure. For some of these, and other, psychiatric disorders there are currently no well-powered GWASs available focussed on clinical diagnostic information. Once these are available, a more comprehensive and accurate underlying structure could be identified that may indeed have important implications for psychiatric nosology. Until then, given the instability of the underlying genetic structure, the identified structure by Lee et al. and those identified in the current study, as well as the inferred implications for psychiatric nosology should be interpreted with caution.

In conclusion, our study illustrated that alcohol and nicotine dependence show widespread genetic correlations with other psychiatric disorders. Including substance use disorders in a genetic factor analysis results in changes in the underlying genetic factor structure of psychiatric disorders. Our study highlights the sensitivity of such (factor) models to the input data. Given the instability of such models, identified structures as well as the implications for psychiatric nosology should be interpreted with caution. To better estimate the most comprehensive and accurate underlying genetic structure, future studies should include data on the broadest possible set of psychiatric disorders.

## Data Availability

All analyses are done on publicly available GWAS summary statistics.

https://www.med.unc.edu/pgc/download-results/

## Acknowledgements

A.A. & K.J.H.V. are supported by the Foundation Volksbond Rotterdam. A.A. is supported by ZonMw grant 849200011 from The Netherlands Organisation for Health Research and Development.

